# Risk of SARS-CoV-2 infection and hospitalization in individuals with natural, vaccine-induced and hybrid immunity: a retrospective population-based cohort study from Estonia

**DOI:** 10.1101/2023.07.18.23292858

**Authors:** Anneli Uusküla, Heti Pisarev, Anna Tisler, Tatjana Meister, Kadri Suija, Kristi Huik, Aare Abroi, Ruth Kalda, Raivo Kolde, Krista Fischer

**Author notes:** **Corresponding Author:** Anneli Uusküla PhD, Department of Family Medicine and Public Health University of Tartu, Tartu 50411, Estonia.

## Abstract

A large proportion of the world’s population has some form of immunity against SARS-CoV- 2, through either infection (‘natural’), vaccination or both (‘hybrid’). This retrospective cohort study used data on SARS-CoV-2, vaccination, and hospitalization from national health system from February 2020 to June 2022 and Cox regression modelling to compare those with natural immunity to those with no (Cohort1, n=92917), hybrid (Cohort2, n=46813), and vaccine (Cohort3, n=252414) immunity. In Cohort 1, those with natural immunity were at lower risk for infection during the Delta (aHR 0.17, 95%CI 0.15-0.18) and higher risk (aHR 1.24, 95%CI 1.18-1.32) during the Omicron period than those with no immunity. Natural immunity conferred substantial protection against COVID-19-hospitalization. Cohort 2 - in comparison to natural immunity hybrid immunity offered strong protection during the Delta (aHR 0.61, 95%CI 0.46-0.80) but not the Omicron (aHR 1.05, 95%CI 0.93-1.1) period. COVID-19-hospitalization was extremely rare among individuals with hybrid immunity. In Cohort 3, individuals with vaccine-induced immunity were at higher risk than those with natural immunity for infection (Delta aHR 4.90, 95%CI 4.48-5.36; Omicron 1.13, 95%CI 1.06-1.21) and hospitalization (Delta aHR 7.19, 95%CI 4.02-12.84). These results show that risk of infection and severe COVID-19 are driven by personal immunity history and the variant of SARS-CoV-2 causing infection.

## Introduction

Despite the large-scale vaccination programs against coronavirus SARS-CoV-2 deployed by governments and health authorities, COVID-19 is continuing to spread.[1] Seroprevalence surveys suggest that more than half of the global population had been infected with SARS-CoV-2 by 2022.[2] Currently, close to three years into the pandemic, a large proportion of the world’s population is likely to have some form of immunity against SARS-CoV-2, either through infection (‘natural’), vaccination (‘artificial’) or both (‘hybrid’). As new variants of SARS-CoV-2 emerge, people are still at risk of new infections and severe COVID-19.

Systematic reviews conducted in the pre-Omicron era agree that vaccines are effective in preventing COVID-19 infection. Studies on the effectiveness of COVID-19 vaccines suggest that protection against SARS-CoV-2 decreases over time, waning considerably after six months. [3,4] There is a large body of evidence documenting that naturally acquired immunity offers strong protection from reinfection. [5,6] Understandably, there is growing interest in comparing the impact of pre-existing SARS-CoV-2 infection-induced immunity [7,8] against hybrid and vaccine-induced immunities.

In several studies, but not all, individuals who had recovered from COVID-19 and were vaccine recipients (BNT162b2, [9–12] mRNA-1273, [10,11] ChAdOx1 [12]) had a significantly lower risk of new SARS-CoV-2 infection than vaccine recipients with no COVID-19 history.[13] Previous SARS-CoV-2 infection and hybrid immunity both provide substantial and sustained protection against Omicron variant.[14] Further, a systematic review demonstrated that natural immunity in COVID-recovered individuals is, at least, equivalent to the protection afforded by complete vaccination of COVID-naïve populations.[15]

However, there are gaps in the literature on the magnitude and durability of protection conferred by different types of SARS-CoV-2 specific immunity when the predominant circulating SARS-CoV-2 strain changes.

We aimed to evaluate the real-world effectiveness of natural immunity in comparison to that of hybrid, vaccine-induced and no immunity against SARS-CoV-2 infection (breakthrough or reinfection) and COVID-19-related hospital admissions and evaluate epidemics stemming from Delta and Omicron SARS-CoV-2 variants in a population-based sample from Estonia.

## Results

### Study population and descriptive statistics

Our analysis was based on data from 246113 matched individuals. Figure 1 shows the dynamic inclusion of individuals in the cohorts over time. The demographic and clinical characteristics of the cohorts are summarized in Table 1. The sex distribution was similar in Cohorts 1 and 2 (with approximately 9.4% more women than men); in Cohort 3, the difference was more pronounced (13.4% more women). The mean age at baseline was 46.2 years in Cohort 1 and 49.3 and 54.3 years in Cohorts 2 and 3, respectively.

**Figure 1.**
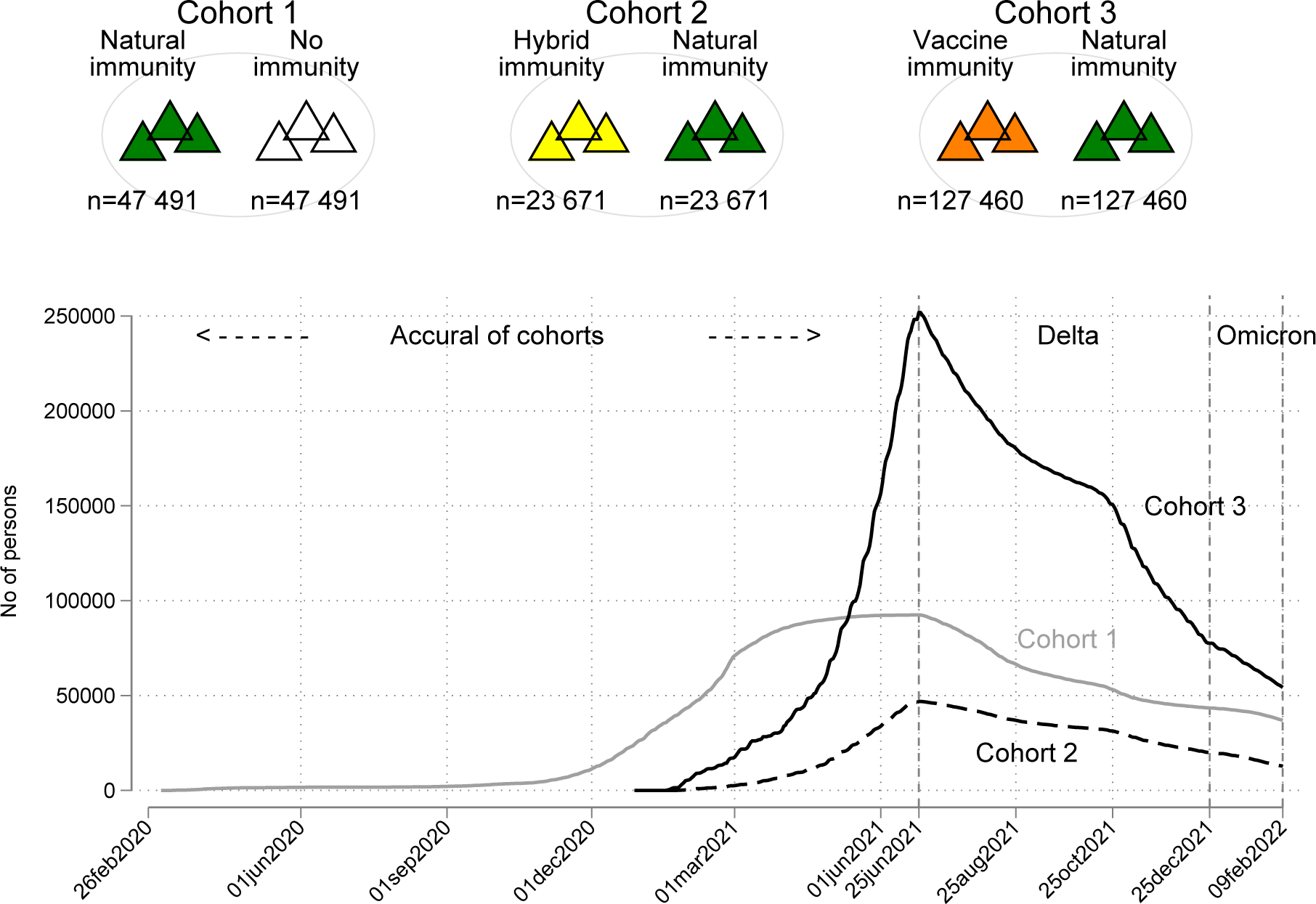
Study cohorts and inclusion to the cohorts.

**Table 1.** Characteristics of study cohorts, baseline dates and follow up times.

| Immunity (sub-cohorts) | Cohort 1 |  | Cohort 2 |  | Cohort 3 |  |
| --- | --- | --- | --- | --- | --- | --- |
|  | Natural | No-immunity | Hybrid | Natural | Vaccine | Natural |
| Number of individuals | 45 820 | 47 097 | 23 567 | 23 462 | 126964 | 125 450 |
| SOCIO-DEMOGRAPHIC CHARACTERISTICS |  |  |  |  |  |  |
| Age, years (mean, SD) | 46 (17) | 47 (17) | 49 (17) | 49 (17) | 54 (18) | 54 (18) |
| Age group (%) |  |  |  |  |  |  |
| 18–49 | 60.8 | 59.2 | 51.6 | 51.8 | 40.6 | 41.1 |
| 50–64 | 24.5 | 24.1 | 28.9 | 29.1 | 26.8 | 27.2 |
| 65–79 | 11.0 | 11.6 | 14.3 | 14.1 | 23.9 | 23.7 |
| 80+ | 3.6 | 5.1 | 5.2 | 5.0 | 8.7 | 8.1 |
| Sex |  |  |  |  |  |  |
| male (%) | 45.4 | 45.4 | 45.3 | 45.4 | 43.3 | 43.3 |
| female (%) | 54.6 | 54.6 | 54.7 | 54.6 | 56.7 | 56.7 |
| Education (%)* |  |  |  |  |  |  |
| below higher | 70.5 | 72.3 | 57.9 | 70.5 | 59.7 | 71.4 |
| higher | 26.7 | 21.3 | 38.5 | 26.2 | 35.3 | 24.6 |
| unknown | 2.9 | 6.4 | 3.7 | 3.3 | 5.0 | 3.9 |
| Nationality (%)† |  |  |  |  |  |  |
| Estonian | 43.3 | 52.7 | 66.5 | 41.6 | 78.6 | 39.1 |
| other | 54.7 | 42.6 | 32.2 | 56.7 | 20.4 | 59.6 |
| unknown |  |  |  |  |  |  |
| PRE-EXISTING COMORBIDITIES |  |  |  |  |  |  |
| Charlson score (%) |  |  |  |  |  |  |
| 0 | 86.4 | 90.2 | 82.4 | 82.5 | 81.0 | 76.5 |
| 1–2 | 10.8 | 8.0 | 13.8 | 13.1 | 15.4 | 17.1 |
| 3+ | 2.8 | 1.8 | 3.8 | 4.4 | 3.5 | 6.5 |
| COVID-19 TESTING AND VACCINATION |  |  |  |  |  |  |
| Intensity of testing per 100 person-day (%) |  |  |  |  |  |  |
| <1 | 79.5 | 87.4 | 89.9 | 85.9 | 90.6 | 95.6 |
| 1... <2 | 16.0 | 9.2 | 7.5 | 9.2 | 5.8 | 3.0 |
| 2+ | 4.5 | 3.5 | 2.6 | 4.9 | 3.5 | 1.5 |
| Number of vaccine doses (% (n)) |  |  |  |  |  |  |
| 1 vaccine dose | 99.1 |  | 83.9 |  |  | 99.6 |
| 2+ vaccine dose | 0.9 |  | 16.1 |  |  | 0.4 |
| COHORT CHARACTERISTICS |  |  |  |  |  |  |
| Baseline date (median, IQR range) | 06feb2021 | 05feb2021 | 14may2021 | 14may2021 | 24may2021 | 25may2021 |
|  | (28dec2020-01mar2021) | (27dec2020-28feb2021) | (21apr2021-03jun2021) | (21apr2021-03jun2021) | (29apr2021-09jun2021) | (29apr2021-09jun2021) |
| Total person-time at risk (days) | 5 807 764 | 7 553 920 | 3 977 206 | 2 938 238 | 16 466 337 | 15 935 771 |
| Total person-time at risk (mean, SD, days) | 127 (85) | 160 (82) | 169 (60) | 125 (85) | 130 (73) | 127 (84) |

### Risk of SARS-CoV-2 infection and COVID-19 hospitalization for individuals with natural immunity compared with individuals with no immunity (Cohort 1)

During the Delta period, the SARS-CoV-2 IR per 100 was 3.8 (95CI 3.5-4.1) for those with natural immunity and over five times higher (IR 20.1 (19.5-20.8)) for those with no immunity. Table 2 shows the number of events (confirmed SARS-CoV-2 infections and hospitalizations) according to the Cohort 1 subcohorts (Table S1 provides a more detailed tabulation of the data by subcohort).

**Table 2.**
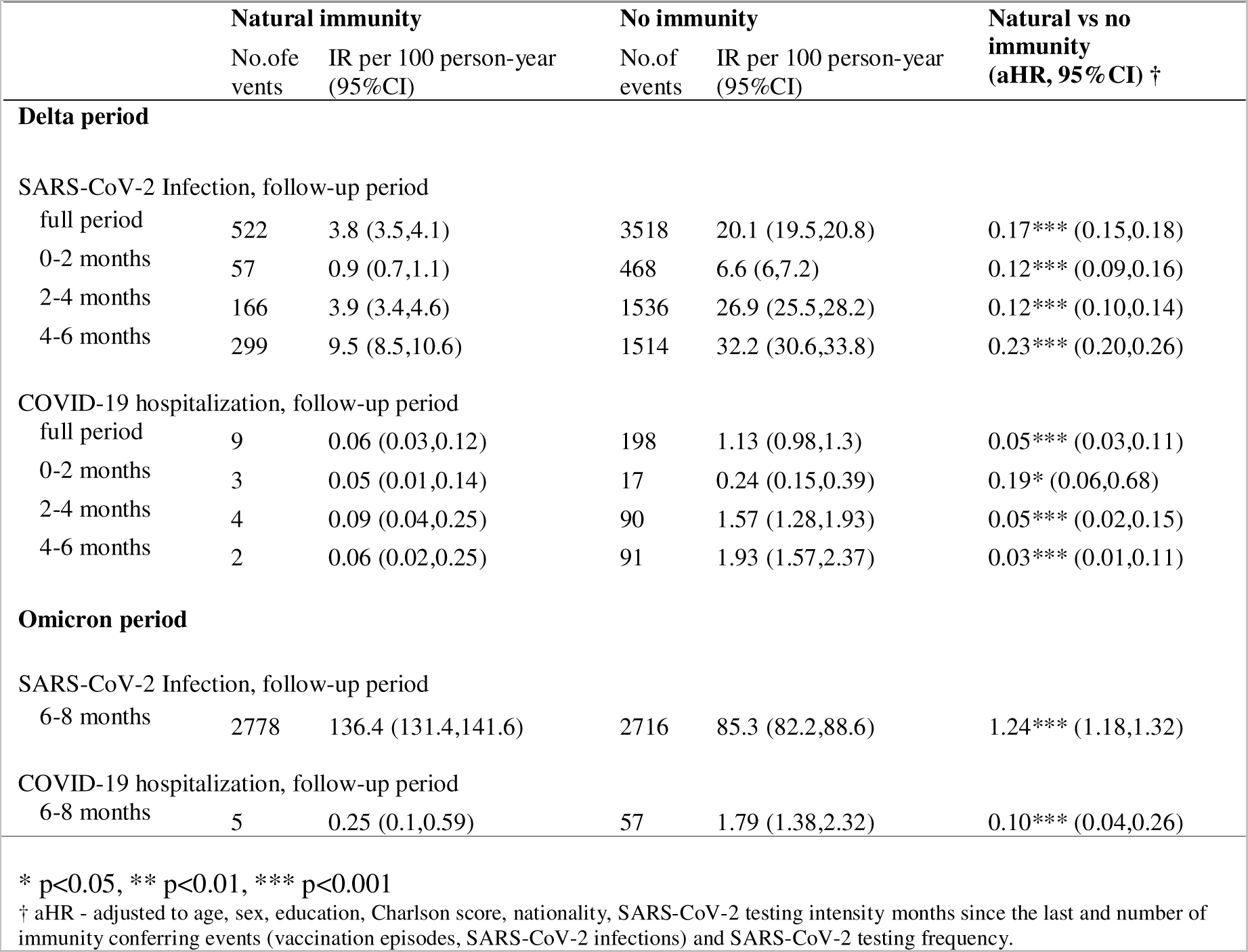
Risk of SARS-CoV-2 infection and COVID-19 hospitalization for individuals with natural immunity compared with individuals with no immunity (Cohort 1)

Figure 2.1 reports the cumulative probability of infection for the Delta (Panel B) and Omicron (Panel C) periods for individuals with natural vs. no immunity.

**Figure 2.**
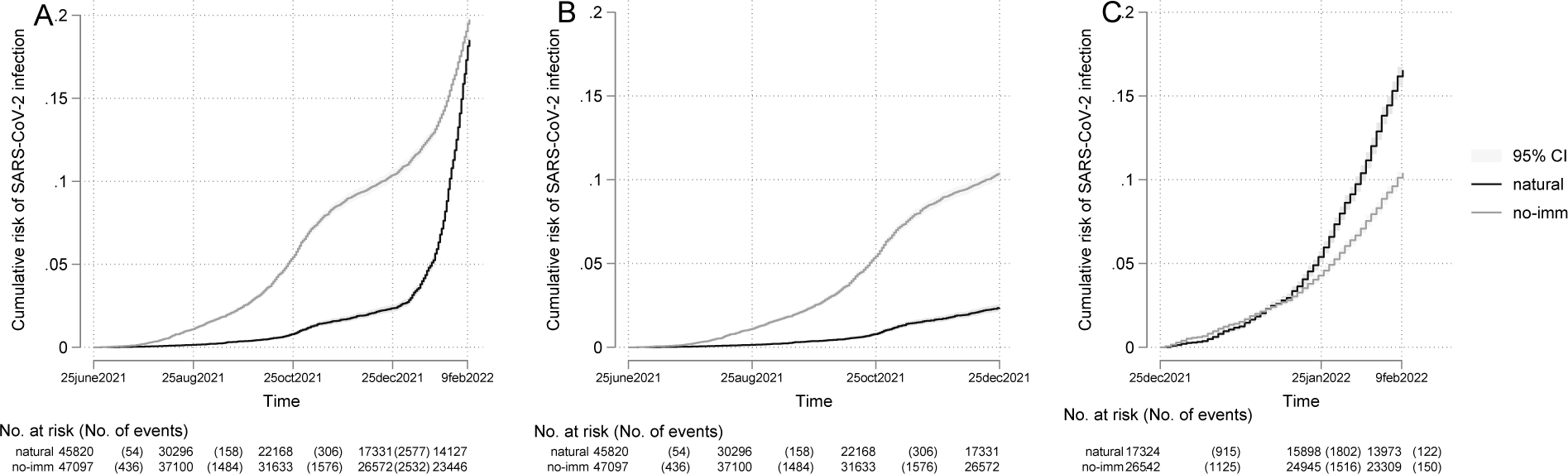
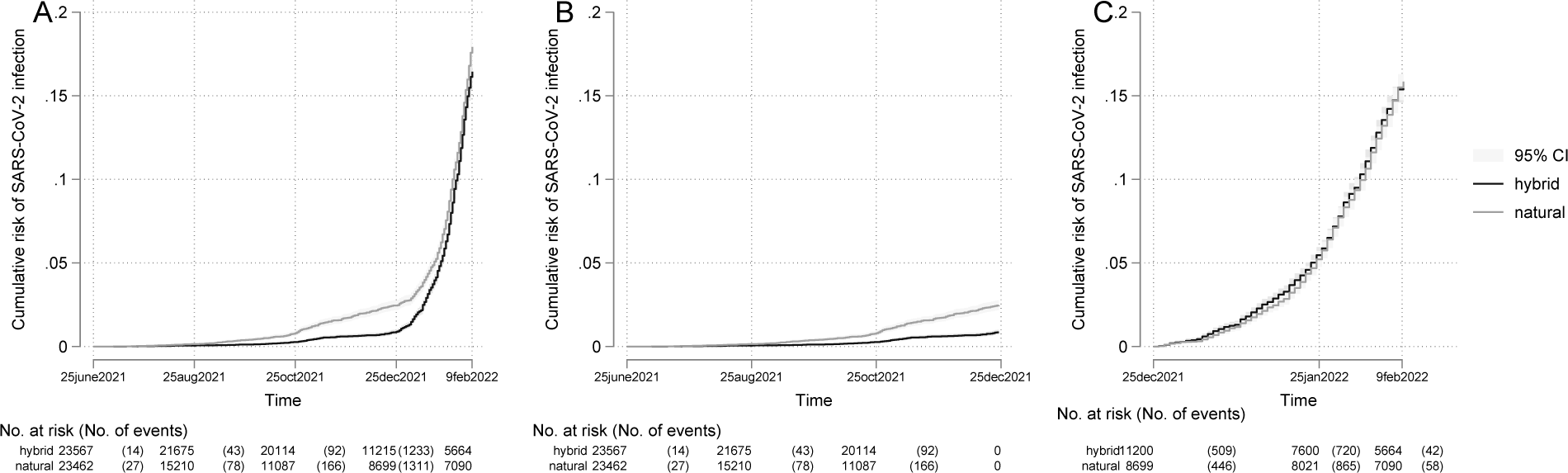
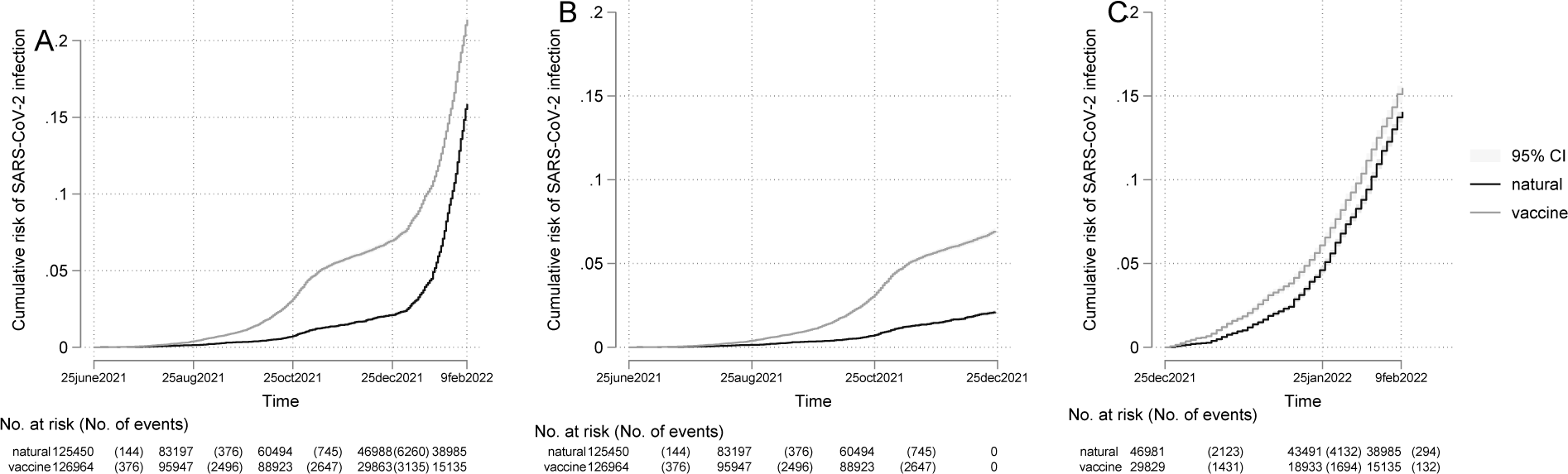
Cumulative risk of infection with SARS-CoV-2, according to cohort and period (overall, Delta, Omicron) Figure 2.1. Cohort 1 - natural and no-immunity (Panels: A overall follow-up period; B Delta period; C Omicron period) Figure 2.2. Cohort 2 - hybrid and natural immunity (Panels: A overall follow-up period; B Delta period; C Omicron period) Figure 2.3. Cohort 3 – natural and vaccine immunity (Panels: A overall follow-up period; B Delta period; C Omicron period)

For this cohort, we observed a reversal of the natural immunity effect. During the whole Delta period, compared to no immunity, natural immunity was associated with a strong and stable protective effect (83%, aHR 0.17, 95%CI 0.15-0.18), but during the Omicron period, there was a 24% higher risk of infection (aHR 1.24, 95%CI 1.18-1.32) (Table 2).

During both periods, natural immunity conferred substantial protection against COVID-19 hospitalization (Delta period aHR 0.05, 95%CI 0.03-0.11); Omicron period aHR 0.10, 95%CI 0.04-0.26) compared to those without SARS-CoV-2-specific immunity.

During both periods (Delta and Omicron), the risk for infection was higher for women (Delta aHR 1.21, 95%CI 1.14-1.29); Omicron aHR 1.1, 95%CI 1.12-1.25), those with more comorbidities, and those younger than 50 years. A longer time since the immunity-conferring event (SARS-CoV-2 infection) was associated with a minimal increase in risk (Delta aHR 1.03, 95%CI 1.01-1.04; Omicron aHR 1.02, 95%CI 1.00-1.03). The complete set of parameter estimates of the regression model is provided in Supplemental Table S4. Increasing age was a significant contributor to the risk for COVID-19 hospitalization, and females had approximately two times lower risk for severe COVID-19 (in comparison to men; Delta aHR 0.59, 95%CI 0.44-0.77, Omicron 0.43, 95%CI 0.26-0.72) (Table S5).

### Risk of SARS-CoV-2 infection and COVID-19 hospitalization for individuals with hybrid immunity compared with individuals with natural immunity (Cohort 2)

During the Delta period, those with hybrid immunity had substantially lower rates of reinfection than those with natural immunity: IR 1.6, 95%CI 1.3-1.8 vs. 3.9, 95%CI 3.4-4.3, but during the Omicron period the difference in risks was diminished (IR 115.8, 95%CI 109.5-122.3 vs. 130.5, 95%CI 123.7-137.7) (see Figure 2.2 and Table 2). During the Delta period, the hybrid immunity subcohort was associated with 39% (aHR 0.61, 95%CI 0.46-0.80) lower risk of infection than the natural immunity subcohort, but this benefit was not sustained during the Omicron period (aHR 1.05, 95%CI 0.93-1.19). Hospitalization due to COVID-19 was extremely rare among those with hybrid immunity (no cases during the Delta period and one case during the Omicron period).

There were differences in other factors associated with the risk of infection during the Delta and Omicron periods (Table S4). During both periods, individuals younger than 50 years had a higher risk for infection. In Delta period, a longer time since the immunity-conferring event was associated with some increase in risk (aHR 1.09, 95%CI 1.05-1.14), as was increasing number of comorbidities (1-2 vs. 0: aHR 1.54, 95%CI 1.16-2.06), 3+ vs 0: 1.38, 95%CI 0.85-2.26). (See Supplemental Tables S2 and S4).

As within Cohort 1, we saw a reversal of the effects of age and sex for cases of COVID-19 infection: increasing age and male sex contributed to the higher risk for severe COVID-19 (Table S5).

### Risk of SARS-CoV-2 infection and COVID-19 hospitalization for individuals with vaccine-induced immunity compared with individuals with natural immunity (Cohort 3)

During the Delta period, individuals with vaccine-induced immunity sustained a higher risk for infection than those with natural immunity (Delta period: IR 13.1, 95%CI 12.7-13.4 vs 3.3, 95%CI 3.2-3.5). For the Omicron period, the difference in risk diminished (IR 116.6, 95%CI 112.6-120.7 vs 115.0, 95%CI 112.2-117.9). The risks for COVID-19 hospitalization were very low in both subcohorts. (See Figure 2.3 and Table 2)

Compared to individuals with natural immunity, those with vaccine-induced immunity were at significantly higher risk of SARS-CoV-2 infection during the Delta period (aHR 4.90, 95%CI 4.48-5.36)), and at 13% higher risk during the Omicron period (aHR 1.13, 95%CI 1.06-1.21). Increased risk for COVID-19 hospitalisation among those with vaccine immunity during the Delta period (aHR 7.19, 95%CI 4.02-12.84) was not sustained over the Omicron period (aHR 2.0, 95%CI 0.64-6.25).

In both periods, the risk of infection was higher among those younger than 50 years, and a longer time since the immunity-conferring event (vaccination) was associated with a minimal increase in risk (Delta 1.13, 95%CI 1.12-1.15; Omicron aHR 1.01, 95%CI 1.00-1.02). (Table S4). The risk of COVID-19 hospitalization was lower among women and increased with age and with increasing numbers of comorbid conditions (Table S5).

## Discussion

This population-based cohort study included data from the national electronic eHealth system to estimate the protection afforded by previous infection, hybrid immunity and vaccine-induced immunity and evaluate epidemics stemming from the Delta and Omicron variants. We observed significant differences in protection against SARS-CoV-2 infection and severe COVID-19 associated with the type of SARS-CoV-2-specific immunity and during the epidemic periods driven by different SARS-CoV-2 variants.

In agreement with other recent studies,[23,24,25] we showed that natural immunity conferred a clear protective effect against infection and hospitalization for more than one year during the Delta variant-dominated period when compared to no SARS-CoV-2-specific immunity.

However, this protective effect against infection was reversed into an increased risk (25%) during the Omicron-dominated period. The decreased effectiveness of natural immunity in preventing reinfection by the new SARS-CoV-2 variant [Omicron] has been described before.^24^ This might be explained by important changes in the virus (Omicron variant being more transmissible and less virulent [26]) and differences in population characteristics (age, contact patterns). During both periods, natural immunity proved to be highly effective in protecting against reinfections progressing to severe disease and was associated with a significantly lower risk of COVID hospitalization than no SARS-CoV-2-specific immunity.

The risk for infection among those with vaccine-induced immunity (in comparison with those with natural immunity) was more pronounced during the Delta variant-dominated period (risk increase of 195% vs. 13% during the Omicron period). The risk of severe COVID-19 among vaccinees in our study is comparable to that described from a UK cohort study (from a similar time period).[27] We were able to extend the results of Gazit et al.[28] by documenting a higher risk for infection and hospitalization caused not only by the Delta variant but also the Omicron SARS-CoV-2 variant among those with vaccine-induced immunity compared to those with natural immunity.

A higher infection protection effect of hybrid immunity in comparison to natural immunity has been described in pre-Omicron studies [11,29] in comparison to natural immunity. During the Delta-dominated period, hybrid immunity offered greater protection against new and severe infections than natural immunity. This effect was not sustained during the Omicron period. Irrespective of the infection-causing variant, the protective effect of hybrid immunity in preventing infection progression to severe COVID-19 significantly exceeded that of natural immunity (although the absolute numbers of hospitalizations in the hybrid immunity subcohort were small).

Our analysis depicts the real-world effectiveness of different immunity-conferring events, and some aspects of our source data need to be discussed. First, our cohorts were accrued over different times (Cohort 1 was the earliest) during the study period. The cohort accrual follows the course of the epidemic and response to the epidemic – from emergence of people with no or post-infection natural immunity (our Cohort 1), progressing to cohorts of individuals with vaccine-induced and hybrid immunities. There are some age differences in cohorts, most likely reflecting the coverage and targeting of epidemic control measures (vaccination; social distancing). Cohort 3 (vaccine-induced vs. natural) members were the oldest (mean 55 years), and Cohort 1 (natural vs. no) members were the youngest (mean 47 years). We also observed some sociodemographic (education) and health/behaviour (vaccination coverage) differences between subcohorts. The majority (84%) of the hybrid immunity subcohort had only one vaccination– this is to be expected as per the vaccination recommendations. However, in the vaccinated subcohort, the majority (73%) had received two vaccine doses (Estonian population coverage with two doses was 62% as of January 2022).[29]

There are important limitations to discuss in our work. First, there is misclassification. Our infected cohort and new infection cohort excluded those who may have had an infection with SARS-CoV-2 but were not tested. The effect of this bias is conservative, leading to underestimation of the true effects. Importantly, the analysis of severe COVID-19 is unlikely to be affected by this bias, as people with symptoms indicative of severe acute respiratory disease would be hospitalized (and tested). Some patients might be admitted to the hospital with, but not because of, SARS-CoV-2 infection. We tried to minimize this by limiting hospitalized COVID-19 cases with ICD-10 diagnoses indicative of respiratory (infection) disease.

Categorization into epidemics driven by two different variants of SARS-CoV-2 was based on calendar time, not on actual test results. Based on the national SARS-CoV-2 genetic surveillance data, we identified periods when variants of concern represented the overwhelming majority of the isolates. Even though in the analysis, we accounted for sex, education, comorbidities, SARS-CoV-2 testing probability and SARS-CoV-2 environmental exposure risk, residual confounding is possible. Our results might be affected by differences between the groups in terms of health behaviours (such as social distancing and mask wearing), a possible confounder that was not assessed. While these biases might influence our estimates, they seem unlikely to have caused the clear patterns observed in this study.

The strengths of this work should be noted. To the best of our knowledge, this is the first study to characterize the risk of infection by more than two different types of SARS-CoV-2-specific immunity using real-life nationwide data. Pairwise comparisons of different immunity states have been reported before.[11,27,29,30] The strength of this work stems from nesting comparator states in the same source population and implementing standardized modelling for effect estimations. Data originating from electronic health and test records are most likely free from recall and social desirability biases. We would like to highlight that our use of a large, population-based sample size increases the generalizability to other countries with similar population structures (and public/health care provisions).

## Conclusions

Our findings suggest that the risk of infection (and of developing severe disease) is affected not only by age and comorbidities but also by personal history of immunity-conferring events and by the viral variant responsible for the epidemic. Therefore, personalized risk-based vaccination strategies could be both effective and cost-effective.

## Methods

### Study setting

Vaccination in Estonia began in January 2021, with a cumulative vaccination uptake about 70% among adult population by June 2022. Within the time period of data underlying the present study, Estonia had three large pandemic waves: the first was from March to June 2020 (SARS-CoV-2 prevariant of concern era); the second was from November 2020 to May 2021 (first the Alpha variant, then the Delta variant); and the third was from December 2022 (Omicron variant).[24] Our analysis used data derived from the nationwide and population-based universal tax-funded Estonian health care system.

### Study design

We conducted a retrospective cohort study based on linking individual-level data on laboratory-confirmed COVID-19 s, SARS-CoV-2 vaccination status, and health care utilization between 26 February 2020 and 23 February 2022 from the national e-health records.[25]

### Data sources

#### The Health and Welfare Information Systems Centre (TEHIK)

Data on COVID-19 vaccination (dates), SARS-CoV-2 testing (dates) and laboratory confirmed (real-time polymerase chain reaction (PCR) or antigen testing) cases of SARS-CoV-2 infections (dates) were retrieved from TEHIK.[26] According to law, all health-care providers and laboratories in Estonia are obligated to report to TEHIK, with an expected coverage of 100%.

#### The Estonian Health Insurance Fund (HIF)

By the end of 2021, universal public health insurance covered 95.2% of the Estonian population (1 328 889 people).[27] The HIF maintains a complete record of the health care services provided. Diagnoses are defined according to the International Classification of Diseases, tenth revision (ICD-10). The HIF database records sex, age, health care utilization information (dates of service, diagnoses, treatment type: in- or outpatient), and date of death.

*The Population Register* is a unified database of Estonian citizens and foreign nationals living in Estonia based on right of residence or residence permits. Population Register data were used to identify the study subjects’ education and ethnicity.

The databases are linked using a unique personal code given to all persons living in Estonia.

### Participants

The source population for this analysis consisted of 343 501 individuals aged 18 years or older. Based on various histories of immunity-conferring events (i.e., infection and/or vaccination) from 26 February 2020 to 25 June 2021, we determined four exposure states:

(i) Individuals with no immunity (SARS-CoV-2 immune-naïve) were defined as those who were unvaccinated and did not have documented previous SARS-CoV-2 infections (n = 130 874);
(ii) Individuals with natural immunity (the recovered, unvaccinated cohort) were those with a documented previous infection but without previous vaccination (n = 47 491);
(iii) Individuals with vaccine-induced SARS-CoV-2 immunity (vaccinated-only cohort) were those without previously recorded infections who received a full vaccination course (BNT162b2; mRNA-1273; AZD1222; Ad26.COV2. S) (n = 127 460); and
(iv) Individuals with hybrid SARS-CoV-2 immunity (the recovered, vaccinated cohort) were defined as those with documented previous infections who received at least one vaccine dose (n = 23 671).

#### Construction of study cohorts

We constructed three mutually exclusive cohorts to assess the risk of SARS-CoV-2 (re)infection and COVID-19 hospital admission. Each cohort consisted of two subcohorts with different types of SARS-CoV-2 immunity (see Figure 1).

Cohort 1 was formed to compare people with natural SARS-CoV-2 immunity to those without SARS-CoV-2 immunity. All individuals with natural immunity were randomly matched (1:1, without replacement) by birth year and sex to unvaccinated individuals with no immunity at baseline (for this cohort, the date of the positive SARS-CoV-2 test for individuals with natural immunity).

Cohort 2 was formed to compare hybrid SARS-CoV-2 immunity with natural immunity. Hybrid immunity was defined as having a documented previous infection and a single vaccine dose either before or after infection or having received two or more vaccine doses, with at least the second dose given after infection (One individual had received three vaccine doses).

All individuals with hybrid immunity were matched to those with natural immunity in a 1:1 ratio (with replacement) based on sex and birth year. For this cohort, the baseline date was defined as (the date of the last immunity-conferring event for individuals with hybrid immunity. Matched subjects with natural immunity had to be alive, previous infected and unvaccinated on the baseline date. Matching was performed as an iterative process until all subjects with hybrid immunity had a suitable match of individuals with natural immunity only. The follow-up started on the baseline date for both individuals in the matched pair.

Cohort 3 was formed to compare vaccine-induced immunity (vaccine only) to natural immunity. Those with vaccine-induced immunity had received at least one vaccine dose (30 individuals had received 3 or more vaccine doses). All individuals with vaccine-induced immunity were matched to those with natural immunity using the same principles as in the second cohort. The follow-up started at baseline (the date of the last immunity-conferring event of the individual with vaccine immunity) for both individuals in the matched pair.

### Study outcomes

The primary outcome was laboratory confirmed SARS-CoV-2 infection occurring after the baseline date: (i) at any time for individuals with no immunity; (ii) after 60 days of recovery from a previous infection for individuals with natural immunity (i.e., reinfection);[18] (iii) after being vaccinated for at least 14 days for individuals with vaccine-induced immunity (SARS-CoV-2 vaccination only) (i.e., breakthrough infection); and (iii) after being vaccinated for at least 14 days or after 60 days [28] of recovery from a previous infection, whichever came later, for individuals with hybrid immunity.

The second outcome was hospitalization with COVID-19 as the reason for admission. This was defined as SARS-CoV-2-related hospitalization occurring from 3 days before to 14 days after a positive SARS-CoV-2 test and the presence of at least one of the following diagnoses (ICD-10) in relation to hospitalization: U07.1, U07.2, acute respiratory tract infections (J00– J06, J12, J15-J18, J20-J22, J46) or severe complications of lower respiratory tract infections (J80–84, J85–J86).[29]

The follow-up duration was counted in days until the date of an outcome, vaccination (for individuals with natural or no immunity), death, or end of the study period (23 February 2022), whichever occurred first.

### Variables accounted for in the risk model

The number of SARS-CoV-2 tests a person received throughout the pandemic was accounted for by counting the number of tests that an individual underwent from baseline to the end of the study. We defined three individualized testing intensities (<1, 1-1.99, ≥2 per 100 person-days).

The comorbidity status was computed based on health data within 12 months before the baseline date using the Charlson comorbidity index (CCI),[30] and study subjects were divided into three groups comprising those with no (CCI score of 0), one or two (CCI score of 1 or 2) or at least three (CCI score ≥3) comorbid conditions.

The follow-up period from baseline was split into four segments: up to 2, 2-4, 4-6, and 6-8 months. The number of and the time since the last immunity-conferring event (since SARS-CoV-2 infection or vaccination) within these groups were also quantified.

### Data analysis

We analysed data from two time periods: from the start of follow-up until 19 December 2021, when the Delta variant was the predominant circulating SARS-CoV-2 strain (proportion of sequenced strains 93%); and 20 December 2021 to 23 February 2022 (end of follow-up), when the Omicron variants (BA1, BA2 and their sublineages) were the predominant strains (proportion of sequenced strains 88%).[31]

Frequencies and proportions for categorical variables, means and standard deviations (SD) for age, and median and range for baseline date were used to characterize the study cohorts (Table 1). The follow-up duration is presented in months. The number of confirmed infections and the crude incidence rates (IRs) per 100 person-years were counted for each cohort (Table S1-S3). Cumulative Kaplan[Meier curves are presented to describe SARS-CoV-2 infections in cohorts by different subcohorts (Figure 2).

We performed Cox regression with the SARS-CoV-2 infection or COVID-19 hospitalization as the dependent variable and sex, age group (18-49, 50-64, 65-79, 80+ years), education (higher, < higher), nationality (Estonian, other), CCI score, time (in months) since the last conferring event and number of conferring events, and SARS-CoV-2 testing intensity as independent variables. Multivariable-adjusted hazard ratios (aHRs) and their 95% confidence intervals (CIs) are presented (Tables 2-4). (See Supplement for additional information on data analysis).

**Table 3.** Risk of SARS-CoV-2 infection and COVID-19 hospitalization for individuals with hybrid immunity compared with individuals with natural immunity (Cohort 2)

|  | Hybrid immunity |  | Natural immunity |  | Hybrid vs natural immunity<br>(aHR, 95%CI) † |
| --- | --- | --- | --- | --- | --- |
|  | No.of events | IR per 100 person-year (95%CI) | No.of events | IR per 100 person-year (95%CI) |  |
| Delta period |  |  |  |  |  |
| SARS-CoV-2 Infection, follow-up period |  |  |  |  |  |
| full period | 153 | 1.6 (1.3,1.8) | 271 | 3.9 (3.4,4.3) | 0.61 *** (0.46,0.80) |
| 0-2 months | 14 | 0.4 (0.2,0.6) | 28 | 0.8 (0.6,1.2) | 0.52 (0.21,1.30) |
| 2-4 months | 43 | 1.2 (0.9,1.7) | 81 | 3.8 (3.1,4.7) | 0.59* (0.36,0.98) |
| 4-6 months | 96 | 3.8 (3.1,4.6) | 162 | 10.2 (8.8,12) | 0.65* (0.45,0.94) |
| COVID-19 hospitalization, follow-up period |  |  |  |  |  |
| full period | - | - | 5 | 0.07 (0.03,0.17) | - |
| Omicron period |  |  |  |  |  |
| SARS-CoV-2 Infection, follow-up period |  |  |  |  |  |
| 6-8 months | 1250 | 115.8 (109.5,122.3) | 1340 | 130.5 (123.7,137.7) | 1.05 (0.93,1.19) |
| COVID-19 hospitalization, follow-up period |  |  |  |  |  |
| 6-8 months | 1 | 0.09 (0.01,0.66) | 4 | 0.39 (0.15,1.04) | 0.70 (0.04,12.70) |

\* p<0.05, \*\* p<0.01, \*\*\* p<0.001
† aHR - adjusted to age, sex, education, Charlson score, nationality, SARS-CoV-2 testing intensity months since the last and number of immunity conferring events (vaccination episodes, SARS-CoV-2 infections) and SARS-CoV-2 testing frequency.

**Table 4.** Risk of SARS-CoV-2 infection and COVID-19 hospitalization for individuals with hybrid immunity compared with individuals with natural immunity (Cohort 3)

|  | Vaccine immunity |  | Natural immunity |  | Vaccine vs natural immunity (aHR, 95%CI) † |
| --- | --- | --- | --- | --- | --- |
|  | No.of events | IR per 100 person-year (95%CI) | No.of events | IR per 100 person-year (95%CI) |  |
| **Delta period** |  |  |  |  |  |
| SARS-CoV-2 Infection, follow-up period |  |  |  |  |  |
| full period | 5529 | 13.1 (12.7,13.4) | 1270 | 3.3 (3.2,3.5) | 4.90*** (4.48,5.36) |
| 0-2 months | 392 | 2.3 (2,2.5) | 147 | 0.8 (0.7,1) | 1.70*** (1.28,2.25) |
| 2-4 months | 2600 | 16.5 (15.9,17.2) | 396 | 3.4 (3.1,3.7) | 6.16*** (5.32,7.14) |
| 4-6 months | 2537 | 27.3 (26.3,28.4) | 727 | 8.5 (7.9,9.1) | 5.10*** (4.48,5.80) |
| COVID-19 hospitalization, follow-up period |  |  |  |  |  |
| full period | 140 | 0.33 (0.28,0.39) | 31 | 0.08 (0.06,0.12) | 7.19*** (4.02,12.84) |
| 0-2 months | 17 | 0.1 (0.06,0.16) | 9 | 0.05 (0.03,0.1) | 1.03 (0.30,3.51) |
| 2-4 months | 69 | 0.44 (0.35,0.56) | 12 | 0.1 (0.06,0.18) | 14.43*** (5.59,37.23) |
| 4-6 months | 54 | 0.58 (0.45,0.76) | 10 | 0.12 (0.06,0.22) | 8.47*** (3.05,23.55) |
| **Omicron period** |  |  |  |  |  |
| SARS-CoV-2 Infection, follow-up period |  |  |  |  |  |
| 6-8 months | 3191 | 116.6 (112.6,120.7) | 6402 | 115.0 (112.2,117.9) | 1.13*** (1.06,1.21) |
| COVID-19 hospitalization, follow-up period |  |  |  |  |  |
| 6-8 months | 14 | 0.51 (0.3,0.8) | 18 | 0.32 (0.2,0.51) | 2.00 (0.64,6.25) |

\* p<0.05, \*\* p<0.01, \*\*\* p<0.001
† aHR - adjusted to age, sex, education, Charlson score, nationality, SARS-CoV-2 testing intensity months since the last and number of immunity conferring events (vaccination episodes, SARS-CoV-2 infections) and SARS-CoV-2 testing frequency.

A p value of less than 0.05 was considered to indicate statistical significance in all analyses. Data analysis was performed with the statistical software Stata 17.0.

The Research Ethics Committee of the University of Tartu approved our study and waived the requirement for informed consent. Whole research was performed in accordance with the relevant guidelines and regulations.

## Author contributions

AU: Conceptualization, Methodology, Data interpretation, Writing original draft; HP: Data curation, Formal analysis, Data interpretation, Visualization, Writing - Review and Editing; KF: Data curation, Formal analysis, Data interpretation, Writing - Review and Editing; AT, TM, AT, KS, RK, KH, RK, KH and AA: Data interpretation, Writing - Review and Editing. All authors contributed to the interpretation of results and critical revision of the manuscript for intellectual content and have given final approval of the version to be published.

## Data availability statement

There are legal restrictions on sharing a de-identified data. According to legislative regulation and data protection law in Estonia, the authors cannot publicly release the data received from the health data registers in Estonia. The data can be requested by completing the application in order to carry out research or an evaluation of public interest (https://www.eithealth-scandinavia.eu/biobanksregisters/access/registers-estonia, and https://www.tehik.ee/en/statistics)

## Competing Interests Statement

The authors declare that they have no competing interests.

## Funding Source

Research was carried out with the support of the European Regional Development Fund (RITA 1/02-120), Estonian Research Council (grants PRG1197, PRG198) and European Social Fund via IT Academy program.

---

**Table 1.** **Characteristics of study cohorts-baseline dates and follow up times.**

| Immunity (sub-cohorts) | Cohort 1 |  | Cohort 2 |  | Cohort 3 |  |
| --- | --- | --- | --- | --- | --- | --- |
|  | Natural | No-immunity | Hybrid | Natural | Vaccine | Natural |
| Number of individuals | 45 820 | 47 097 | 23 567 | 23 462 | 126964 | 125 450 |
| SOCIO-DEMOGRAPHIC CHARACTERISTICS |  |  |  |  |  |  |
| Age- years (mean- SD) | 46 (17) | 47 (17) | 49 (17) | 49 (17) | 54 (18) | 54 (18) |
| Age group (%) |  |  |  |  |  |  |
| 18–49 | 60.8 | 59.2 | 51.6 | 51.8 | 40.6 | 41.1 |
| 50–64 | 24.5 | 24.1 | 28.9 | 29.1 | 26.8 | 27.2 |
| 65–79 | 11.0 | 11.6 | 14.3 | 14.1 | 23.9 | 23.7 |
| 80+ | 3.6 | 5.1 | 5.2 | 5.0 | 8.7 | 8.1 |
| Sex |  |  |  |  |  |  |
| male (%) | 45.4 | 45.4 | 45.3 | 45.4 | 43.3 | 43.3 |
| female (%) | 54.6 | 54.6 | 54.7 | 54.6 | 56.7 | 56.7 |
| Education (%)* |  |  |  |  |  |  |
| below higher | 70.5 | 72.3 | 57.9 | 70.5 | 59.7 | 71.4 |
| higher | 26.7 | 21.3 | 38.5 | 26.2 | 35.3 | 24.6 |
| unknown | 2.9 | 6.4 | 3.7 | 3.3 | 5.0 | 3.9 |
| Nationality (%)† |  |  |  |  |  |  |
| Estonian | 43.3 | 52.7 | 66.5 | 41.6 | 78.6 | 39.1 |
| other | 54.7 | 42.6 | 32.2 | 56.7 | 20.4 | 59.6 |
| unknown |  |  |  |  |  |  |
| PRE-EXISTING COMORBIDITIES |  |  |  |  |  |  |
| Charlson score (%) |  |  |  |  |  |  |
| 0 | 86.4 | 90.2 | 82.4 | 82.5 | 81.0 | 76.5 |
| 1–2 | 10.8 | 8.0 | 13.8 | 13.1 | 15.4 | 17.1 |
| 3+ | 2.8 | 1.8 | 3.8 | 4.4 | 3.5 | 6.5 |
| COVID-19 TESTING AND VACCINATION |  |  |  |  |  |  |
| Intensity of testing per 100 person-day (%) |  |  |  |  |  |  |
| <1 | 79.5 | 87.4 | 89.9 | 85.9 | 90.6 | 95.6 |
| 1... <2 | 16.0 | 9.2 | 7.5 | 9.2 | 5.8 | 3.0 |
| 2+ | 4.5 | 3.5 | 2.6 | 4.9 | 3.5 | 1.5 |
| Number of vaccine doses (% (n)) |  |  |  |  |  |  |
| 1 vaccine dose | 99.1 |  | 83.9 |  |  | 99.6 |
| 2+ vaccine dose | 0.9 |  | 16.1 |  |  | 0.4 |
| COHORT CHARACTERISTICS |  |  |  |  |  |  |
| Baseline date (median- IQR range) | 06feb2021<br>(28dec2020-01mar2021) | 05feb2021<br>(27dec2020-28feb2021) | 14may2021<br>(21apr2021-03jun2021) | 14may2021<br>(21apr2021-03jun2021) | 24may2021<br>(29apr2021-09jun2021) | 25may2021<br>(29apr2021-09jun2021) |
| Total person-time at risk (days) | 5 807 764 | 7 553 920 | 3 977 206 | 2 938 238 | 16 466 337 | 15 935 771 |
| Total person-time at risk (mean- SD- days) | 127 (85) | 160 (82) | 169 (60) | 125 (85) | 130 (73) | 127 (84) |

**Table 2.** Risk of SARS-CoV-2 infection and COVID-19 hospitalization for study Cohorts

| **Cohort 1: individuals with natural immunity compared with individuals with no immunity** |  |  |  |  |  |
| --- | --- | --- | --- | --- | --- |
|  | Natural immunity |  | No immunity |  | Natural vs no immunity (aHR- 95%CI) † |
|  | No.of events | IR per 100 person-year (95%CI) | No.of events | IR per 100 person-year (95%CI) |  |
| **Delta period** |  |  |  |  |  |
| SARS-CoV-2 Infection- follow-up period |  |  |  |  |  |
| full period | 522 | 3.8 (3.5-4.1) | 3518 | 20.1 (19.5-20.8) | 0.17*** (0.15-0.18) |
| 0-2 months | 57 | 0.9 (0.7-1.1) | 468 | 6.6 (6.0-7.2) | 0.12*** (0.09-0.16) |
| 2-4 months | 166 | 3.9 (3.4-4.6) | 1536 | 26.9 (25.5-28.2) | 0.12*** (0.10-0.14) |
| 4-6 months | 299 | 9.5 (8.5-10.6) | 1514 | 32.2 (30.6-33.8) | 0.23*** (0.20-0.26) |
| COVID-19 hospitalization- follow-up period |  |  |  |  |  |
| full period | 9 | 0.06 (0.03-0.12) | 198 | 1.13 (0.98-1.3) | 0.05*** (0.03-0.11) |
| 0-2 months | 3 | 0.05 (0.01-0.14) | 17 | 0.24 (0.15-0.39) | 0.19* (0.06-0.68) |
| 2-4 months | 4 | 0.09 (0.04-0.25) | 90 | 1.57 (1.28-1.93) | 0.05*** (0.02-0.15) |
| 4-6 months | 2 | 0.06 (0.02-0.25) | 91 | 1.93 (1.57-2.37) | 0.03*** (0.01-0.11) |
| **Omicron period** |  |  |  |  |  |
| SARS-CoV-2 Infection- follow-up period |  |  |  |  |  |
| 6-8 months | 2778 | 136.4 (131.4-141.6) | 2716 | 85.3 (82.2-88.6) | 1.24*** (1.18-1.32) |
| COVID-19 hospitalization- follow-up period |  |  |  |  |  |
| 6-8 months | 5 | 0.25 (0.1-0.59) | 57 | 1.79 (1.38-2.32) | 0.10*** (0.04-0.26) |
| **Cohort 2: individuals with hybrid immunity compared with individuals with natural immunity** |  |  |  |  |  |
|  | Hybrid immunity |  | Natural immunity |  | Hybrid vs natural immunity (aHR. 95%CI) † |
|  | No.of events | IR per 100 person-year (95%CI) | No.of events | IR per 100 person-year (95%CI) |  |
| **Delta period** |  |  |  |  |  |
| SARS-CoV-2 Infection- follow-up period |  |  |  |  |  |
| full period | 153 | 1.6 (1.3-1.8) | 271 | 3.9 (3.4-4.3) | 0.61*** (0.46-0.80) |
| 0-2 months | 14 | 0.4 (0.2-0.6) | 28 | 0.8 (0.6-1.2) | 0.52 (0.21-1.30) |
| 2-4 months | 43 | 1.2 (0.9-1.7) | 81 | 3.8 (3.1-4.7) | 0.59* (0.36-0.98) |
| 4-6 months | 96 | 3.8 (3.1-4.6) | 162 | 10.2 (8.8-12.0) | 0.65* (0.45-0.94) |
| COVID-19 hospitalization- follow-up period |  |  |  |  |  |
| full period | - | - | 5 | 0.07 (0.03-0.17) | - |
| **Omicron period** |  |  |  |  |  |
| SARS-CoV-2 Infection- follow-up period |  |  |  |  |  |
| 6-8 months | 1250 | 115.8 (109.5-122.3) | 1340 | 130.5 (123.7-137.7) | 1.05 (0.93-1.19) |
| COVID-19 hospitalization- follow-up period |  |  |  |  |  |
| 6-8 months | 1 | 0.09 (0.01-0.66) | 4 | 0.39 (0.15-1.04) | 0.70 (0.04-12.70) |

Cohort 3: individuals with hybrid immunity compared with individuals with natural immunity
|  | Vaccine immunity |  | Natural immunity |  | Vaccine vs natural immunity |
| --- | --- | --- | --- | --- | --- |
|  | No.of events | IR per 100 person-year (95%CI) | No.of events | IR per 100 person-year (95%CI) | (aHR- 95%CI) † |
| Delta period |  |  |  |  |  |
| SARS-CoV-2 Infection- follow-up period |  |  |  |  |  |
| full period | 5529 | 13.1 (12.7-13.4) | 1270 | 3.3 (3.2-3.5) | 4.90*** (4.5-5.4) |
| 0-2 months | 392 | 2.3 (2.0-2.5) | 147 | 0.8 (0.7-1.0) | 1.70*** (1.3-2.3) |
| 2-4 months | 2600 | 16.5 (15.9-17.2) | 396 | 3.4 (3.1-3.7) | 6.16*** (5.3-7.1) |
| 4-6 months | 2537 | 27.3 (26.3-28.4) | 727 | 8.5 (7.9-9.1) | 5.10*** (4.5-5.8) |
| COVID-19 hospitalization- follow-up period |  |  |  |  |  |
| full period | 140 | 0.33 (0.28-0.39) | 31 | 0.08 (0.06-0.12) | 7.19*** (4.0-12.8) |
| 0-2 months | 17 | 0.1 (0.06-0.16) | 9 | 0.05 (0.03-0.10) | 1.03 (0.30-3.51) |
| 2-4 months | 69 | 0.44 (0.35-0.56) | 12 | 0.1 (0.06-0.18) | 14.43*** (5.6-37.2) |
| 4-6 months | 54 | 0.58 (0.45-0.76) | 10 | 0.12 (0.06-0.22) | 8.47*** (3.1-23.6) |
| Omicron period |  |  |  |  |  |
| SARS-CoV-2 Infection- follow-up period |  |  |  |  |  |
| 6-8 months | 3191 | 116.6 (112.6-120.7) | 6402 | 115.0 (112.2-117.9) | 1.13*** (1.06-1.21) |
| COVID-19 hospitalization- follow-up period |  |  |  |  |  |
| 6-8 months | 14 | 0.51 (0.3-0.8) | 18 | 0.32 (0.2-0.51) | 2.00 (0.64-6.25) |

\* p<0.05- \*\* p<0.01- \*\*\* p<0.001
† aHR - adjusted to age- sex- education- Charlson score- nationality- SARS-CoV-2 testing intensity months since the last and number of immunity conferring events (vaccination episodes- SARS-CoV-2 infections) and SARS-CoV-2 testing frequency.

## Notes

### Competing Interest Statement

The authors have declared no competing interest.

### Author Declarations

The Research Ethics Committee of the University of Tartu approved our study and waived the requirement for informed consent.

