## Supplement for "Risk of SARS-CoV-2 infection and hospitalization in individuals with natural, vaccine-induced and hybrid immunity: a retrospective population-based cohort study from Estonia"

### Content

Statistical analysis.

Table S1. Risk of SARS-CoV-2 infection in Delta and Omicron periods, Cohort 1 (natural vs no-immunity)

Table S2. Risk of SARS-CoV-2 infection in Delta end Omicron periods, Cohort 2 (hybrid vs natural immunity)

Table S3. Risk of SARS-CoV-2 infection in Delta end Omicron periods, Cohort 3 (vaccine vs natural immunity)

Table S4. Adjusted HR-sFactors associated with SARS-CoV-2 infection (adjusted HRs together with 95% CI), Estonia 2020–2022.

Table S5. Factors associated with SARS-CoV-2 infection (adjusted HRs together with 95% CI), Estonia 2020–2022.

**Statistical analysis**

We analysed data from two time periods: from the start of follow-up until 19 December 2021, when the Delta variant was the predominant circulating SARS-CoV-2 strain (proportion of sequenced strains 93%); and 20 December 2021 to 23 February 2022 (end of follow-up), when the Omicron variants (BA1, BA2 and their sublineages) were the predominant strains (proportion of sequenced strains 88%).[21]

We used frequencies and proportions for categorical variables, means and standard deviations (SD) for age, and median and range for baseline date to characterize the study cohorts (Table 1). The follow-up duration is presented in months. The number of confirmed infections and the crude incidence rates (IRs) per 100 person-years were counted for each cohort (Table S1-S3). Cumulative Kaplan–Meier curves are presented to describe SARS-CoV-2 infections in cohorts by different subcohorts (Figure 2).

We performed Cox regression with the SARS-CoV-2 infection or COVID-19 hospitalization as the dependent variable and sex, age group (18-49, 50-64, 65-79, 80+ years), education (higher, < higher), nationality (Estonian, other), CCI score, time (in months) since the last conferring event and number of conferring events, and SARS-CoV-2 testing intensity as independent variables. Multivariable-adjusted hazard ratios (aHRs) and their 95% confidence intervals (CIs) are presented (Tables 2-4).

We developed separate regression models using the same set of independent and dependent variables for the Delta period and the Omicron period. Note that the calendar time affects the hazards of the different cohorts only via the time-dependence of the SARS-CoV-2 exposure risk (background infection rate).

To assess change in the risk of outcomes (and to fulfil the hazards proportionality assumption), we used piecewise hazards models, whereby follow-up time was split into segments (with a 2-month duration), and hazard ratios were calculated separately for each time period by applying a full Cox model (as described above) within each time segment (this was done for both the Delta and Omicron periods).

For Cohorts two and three, based on the matching procedure, weights were assigned to individuals proportional to the number of unexposed individuals matched to each exposed subject. The weights were used in Cox regression models.

A p value of less than 0.05 was considered to indicate statistical significance in all analyses.

Data analysis was performed with the statistical software Stata 17.0, and the CCI was calculated using the macro *charlson*.

**Table S1. Risk of SARS-CoV-2 (re)infection in Delta end Omicron periods cohort 1 (natural vs no-immunity)**

| | Delta period | | | | Omicron period | | | |
|---|---|---|---|---|---|---|---|---|
| | **natural** | | **no-imm** | | **natural** | | **no-imm** | |
| | Number of events | IR per 100 person-year (95% CI) | Number of events | IR per 100 person-year (95% CI) | Number of events | IR per 100 person-year (95% CI) | Number of events | IR per 100 person-year (95% CI) |
| Full follow-up period | 520 | 3.8 (3.5,4.1) | 3507 | 20.1 (19.5,20.8) | 2778 | 136.4 (131.4,141.6) | 2716 | 85.3 (82.2,88.6) |
| Hospitalized | 14 | 0.09 (0.05,0.15) | 255 | 1.23 (1.09,1.39) | 5 | 0.25 (0.1,0.59) | 57 | 1.79 (1.38,2.32) |
| Age groups | | | | | | | | |
| 18–49 years | 388 | 4.6 (4.2,5.1) | 2404 | 24.2 (23.2,25.2) | 2075 | 168.2 (161.1,175.6) | 1926 | 109.8 (105,114.8) |
| 50–64 years | 70 | 2.1 (1.7,2.7) | 747 | 17.5 (16.3,18.8) | 536 | 110 (101.1,119.7) | 592 | 74 (68.3,80.2) |
| 65–79 years | 38 | 2.4 (1.8,3.4) | 247 | 11.3 (9.9,12.8) | 143 | 64.3 (54.5,75.7) | 153 | 35.7 (30.5,41.8) |
| 80+ years | 24 | 4.1 (2.7,6.1) | 109 | 10.7 (8.9,13.) | 24 | 25.7 (17.3,38.4) | 45 | 22.5 (16.8,30.2) |
| Sex | | | | | | | | |
| Male | 214 | 3.5 (3.0,4.0) | 1384 | 17.8 (16.8,18.7) | 1183 | 131.9 (124.6,139.7) | 1062 | 74.6 (70.3,79.3) |
| Female | 306 | 4.0 (3.6,4.5) | 2123 | 22 (21.1,23.0) | 1595 | 139.9 (133.2,146.9) | 1654 | 94 (89.6,98.6) |
| Education | | | | | | | | |
| below higher | 393 | 3.9 (3.6,4.3) | 2660 | 21.3 (20.5,22.1) | 2016 | 133.9 (128.2,139.9) | 2050 | 91.8 (87.9,95.8) |
| higher | 115 | 3.4 (2.9,4.1) | 772 | 21.2 (19.8,22.8) | 723 | 158.1 (147,170.1) | 613 | 92.8 (85.7,100.4) |
| unknown | 12 | 2.8 (1.6,4.9) | 75 | 5.8 (4.6,7.2) | 39 | 53.1 (38.8,72.6) | 53 | 18.4 (14,24.1) |
| Nationality | | | | | | | | |
| estonian | 249 | 4.4 (3.0.9,5) | 2227 | 25.2 (24.2,26.2) | 1196 | 147 (138.9,155.5) | 1570 | 99.5 (94.7,104.6) |
| else | 260 | 3.3 (2.0.9,3.7) | 1230 | 16.1 (15.2,17) | 1547 | 132.1 (125.6,138.8) | 1100 | 78.6 (74.1,83.4) |
| unknown | 11 | 3.7 (2,6.6.0) | 50 | 5.4 (4.1,7.1) | 35 | 68 (48.8,94.7) | 46 | 22.3 (16.7,29.8) |
| Charlson score | | | | | | | | |
| 0 | 447 | 3.8 (3.4,4.1) | 3174 | 20.3 (19.6,21) | 2506 | 143.8 (138.3,149.5) | 2481 | 86.4 (83.1,89.9) |
| 1–2 | 57 | 3.7 (2.8,4.7) | 257 | 17.7 (15.7,20.0) | 234 | 100.9 (88.7,114.7) | 214 | 82.9 (72.5,94.8) |
| 3+ | 16 | 3.8 (2.3,6.2) | 76 | 24.1 (19.3,30.2) | 38 | 61.5 (44.7,84.5) | 21 | 39.2 (25.5,60.1) |

| | | | | | | | | |
|---|---|---|---|---|---|---|---|---|
| Intensity of testing per 100 days | | | | | | | | |
| <1 | 225 | 2.1 (1.8,2.4) | 2469 | 16 (15.4,16.6) | 1679 | 105.3 (100.3,110.4) | 1985 | 68.1 (65.2,71.2) |
| 1..<2 | 233 | 10.1 (8.9,11.5) | 779 | 52.7 (49.1,56.5) | 853 | 262.9 (245.9,281.2) | 537 | 263.7 (242.3,287.0) |
| 2+ | 62 | 8.6 (6.7,11.1) | 259 | 50.1 (44.4,56.6) | 246 | 210.3 (185.6,238.3) | 194 | 296.6 (257.7,341.5) |
| Time since the last immunity conferring event | | | | | | | | |
| <100 days | 68 | 2.9 (2.3,3.6) | 530 | 20.3 (18.6,22.1) | 378 | 119.2 (107.7,131.8) | 401 | 84.2 (76.3,92.8) |
| 100-199 days | 339 | 3.6 (3.0.3,4) | 2470 | 20 (19.2,20.8) | 1909 | 138.5 (132.5,144.9) | 1908 | 84.5 (80.8,88.4) |
| ≥200 days | 113 | 5.5 (4.0.6,6.6) | 507 | 20.7 (18.9,22.5) | 491 | 143.8 (131.6,157.1) | 407 | 90.8 (82.4,100) |
| No of immunity conferring events | | | | | | | | |
| 1 | 518 | 3.8 (3.5,4.1) | 3479 | 20.1 (19.5,20.8) | 2728 | 137.2 (132.1,142.4) | 2692 | 85.4 (82.2,88.6) |
| ≥2 | 2 | 1 (0.2,3.9) | 28 | 17.9 (12.3,25.9) | 50 | 104.3 (79.1,137.6) | 24 | 83 (55.7,123.9) |

**Table S2. Risk of SARS-CoV-2 (re)infection in Delta end Omicron periods cohort 2 (hybrid vs natural)**

| | Delta period | | | | Omicron period | | | |
|---|---|---|---|---|---|---|---|---|
| | **hybrid** | | **natural** | | **hybrid** | | **natural** | |
| | Number of events | IR per 100 person-year (95% CI) | Number of events | IR per 100 person-year (95% CI) | Number of events | IR per 100 person-year (95% CI) | Number of events | IR per 100 person-year (95% CI) |
| Full follow-up period | 149 | 1.5 (1.3,1.8) | 271 | 3.9 (3.4,4.4) | 1250 | 115.8 (109.5,122.3) | 1340 | 130.5 (123.7,137.7) |
| Hospitalized | | | | | 1 | 0.09 (0.01,0.66) | 4 | 0.39 (0.15,1.04) |
| Age groups | | | | | | | | |
| 18–49 years | 86 | 1.7 (1.4,2.1) | 173 | 4.8 (4.1,5.6) | 920 | 144.2 (135.2,153.8) | 892 | 168.6 (157.9,180) |
| 50–64 years | 29 | 1 (0.7,1.5) | 53 | 2.7 (2.1,3.5) | 265 | 87.9 (78,99.2) | 339 | 116.3 (104.6,129.4) |
| 65–79 years | 24 | 1.8 (1.2,2.6) | 24 | 2.4 (1.6,3.6) | 45 | 41.7 (31.1,55.8) | 90 | 63.3 (51.5,77.8) |
| 80+ years | 10 | 2.1 (1.1,3.9) | 21 | 5.1 (3.3,7.8) | 20 | 61.6 (39.7,95.4) | 19 | 29.7 (18.9,46.5) |
| Sex | | | | | | | | |
| Male | 62 | 1.4 (1.1,1.8) | 100 | 3.2 (2.6,3.9) | 567 | 113.8 (104.8,123.6) | 575 | 128.3 (118.3,139.3) |
| Female | 87 | 1.6 (1.3,2.0) | 171 | 4.4 (3.8,5.1) | 683 | 117.4 (108.9,126.5) | 765 | 132.2 (123.2,141.9) |
| Education | | | | | | | | |
| below higher | 88 | 1.5 (1.2,1.9) | 206 | 4.1 (3.5,4.6) | 736 | 110.5 (102.8,118.8) | 981 | 129.3 (121.4,137.6) |
| higher | 52 | 1.4 (1.1,1.8) | 57 | 3.4 (2.6,4.5) | 492 | 126.8 (116.1,138.5) | 340 | 150.4 (135.2,167.2) |
| unknown | 9 | 2.7 (1.4,5.1) | 8 | 3.2 (1.6,6.3) | 22 | 85.8 (56.5,130.3) | 19 | 45.7 (29.1,71.6) |
| Nationality | | | | | | | | |
| estonian | 96 | 1.5 (1.2,1.8) | 127 | 4.6 (3.9,5.5) | 790 | 118.9 (110.9,127.4) | 551 | 139.3 (128.1,151.4) |
| else | 52 | 1.6 (1.2,2.1) | 137 | 3.3 (2.8,3.9) | 441 | 110.5 (100.6,121.3) | 775 | 127.3 (118.7,136.6) |
| unknown | 1 | 0.8 (0.1,5.7) | 7 | 5.3 (2.5,11.1) | 19 | 118.1 (75.3,185.1) | 14 | 62.3 (36.9,105.1) |
| Charlson score | | | | | | | | |
| 0 | 113 | 1.4 (1.2,1.7) | 213 | 3.7 (3.3,4.3) | 1127 | 120.5 (113.6,127.7) | 1178 | 140.2 (132.4,148.4) |
| 1–2 | 25 | 1.9 (1.3,2.8) | 47 | 5 (3.7,6.6) | 104 | 86.9 (71.7,105.4) | 135 | 97.2 (82.1,115.0) |
| 3+ | 11 | 3.1 (1.7,5.7) | 11 | 3.3 (1.0.8,6) | 19 | 76.9 (49.1,120.6) | 27 | 56.9 (39.0,83.0) |
| Intensity of testing per 100 days | | | | | | | | |

| | | | | | | | | |
|---|---|---|---|---|---|---|---|---|
| <1 | 90 | 1 (0.8,1.3) | 128 | 2.2 (1.9,2.6) | 931 | 97.2 (91.2,103.7) | 876 | 103.3 (96.7,110.4) |
| 1– <2 | 41 | 5.4 (4,7.4.0) | 86 | 10.9 (8.9,13.5) | 245 | 265.4 (234.1,300.8) | 308 | 255.1 (228.2,285.3) |
| 2+ | 18 | 7.7 (4.8,12.2) | 57 | 14.4 (11.1,18.7) | 74 | 245.4 (195.4,308.1) | 156 | 268.4 (229.4,313.9) |
| Time since the last immunity conferring event | | | | | | | | |
| <100 days | 120 | 1.3 (1.1,1.6) | 27 | 3 (2.1,4.4) | 1183 | 115.5 (109.1,122.3) | 127 | 113.6 (95.4,135.1) |
| 100-199 days | 29 | 3.4 (2.3,4.9) | 180 | 3.6 (3.1,4.2) | 67 | 119.4 (94.0,151.7) | 945 | 129.3 (121.3,137.8) |
| ≥200 days | | | 64 | 5.8 (4.5,7.4) | | | 268 | 145.7 (129.3,164.2) |
| No of immunity conferring events | | | | | | | | |
| 1 | 149 | 1.5 (1.3,1.8) | 270 | 3.9 (3.4,4.4) | 1250 | 115.8 (109.6,122.4) | 1333 | 130.2 (123.4,137.4) |
| ≥2 | | | 1 | 4.9 (0.7,34.7) | | | 7 | 260.9 (124.4,547.3) |

**Table S3. Risk of SARS-CoV-2 (re)infection in Delta end Omicron periods cohort 3 (vaccine vs natural)**

| | Delta period | | | | Omicron period | | | |
|---|---|---|---|---|---|---|---|---|
| | **vaccine** | | **natural** | | **vaccine** | | **natural** | |
| | Number of events | IR per 100 person-year (95% CI) | Number of events | IR per 100 person-year (95% CI) | Number of events | IR per 100 person-year (95% CI) | Number of events | IR per 100 person-year (95% CI) |
| Full follow-up period | 5519 | 13.1 (12.7,13.4) | 1265 | 3.3 (3.2,3.5) | 3191 | 116.6 (112.6,120.7) | 1265 | 3.3 (3.2,3.5) |
| Hospitalized | 140 | 0.33 (0.28,0.39) | 31 | 0.08 (0.06,0.12) | 14 | 0.51 (0.3,0.8) | 18 | 0.32 (0.2,0.51) |
| Age groups | | | | | | | | |
| 18–49 years | 2493 | 18 (17.4,18.8) | 692 | 4.5 (4.2,4.8) | 1903 | 175.9 (168.2,184) | 692 | 4.5 (4.2,4.8) |
| 50–64 years | 1700 | 13.3 (12.7,14) | 221 | 2.2 (2.0,2.6) | 956 | 106.3 (99.8,113.3) | 221 | 2.2 (2.0,2.6) |
| 65–79 years | 948 | 8.2 (7.7,8.8) | 216 | 2.4 (2.1,2.7) | 270 | 48.7 (43.2,54.8) | 216 | 2.4 (2.1,2.7) |
| 80+ years | 378 | 9 (8.2,10.0) | 136 | 3.9 (3.3,4.6) | 62 | 30.8 (24.0,39.5) | 136 | 3.9 (3.3,4.6) |
| Sex | | | | | | | | |
| Male | 2271 | 12.7 (12.2,13.3) | 499 | 3.1 (2.9,3.4) | 1333 | 111.9 (106,118) | 499 | 3.1 (2.9,3.4) |
| Female | 3248 | 13.3 (12.8,13.8) | 766 | 3.5 (3.2,3.7) | 1858 | 120.2 (114.8,125.8) | 766 | 3.5 (3.2,3.7) |
| Education | | | | | | | | |
| below higher | 3068 | 11.9 (11.5,12.3) | 963 | 3.5 (3.2,3.7) | 1981 | 106.3 (101.7,111.1) | 963 | 3.5 (3.2,3.7) |
| higher | 2195 | 15.5 (14.9,16.2) | 252 | 3 (2.6,3.4) | 1144 | 151.9 (143.4,161) | 252 | 3 (2.6,3.4) |
| unknown | 256 | 10.9 (9.7,12.3) | 50 | 3.2 (2.4,4.2) | 66 | 54.4 (42.7,69.2) | 50 | 3.2 (2.4,4.2) |
| Nationality | | | | | | | | |
| estonian | 4492 | 13.6 (13.2,14.0) | 569 | 4.1 (3.8,4.4) | 2333 | 117.9 (113.2,122.7) | 569 | 4.1 (3.8,4.4) |
| else | 978 | 10.9 (10.3,11.6) | 677 | 2.9 (2.7,3.1) | 825 | 113.6 (106.1,121.7) | 677 | 2.9 (2.7,3.1) |
| unknown | 49 | 13.2 (10.0,17.5) | 19 | 3.5 (2.2,5.4) | 33 | 102.7 (73,144.4) | 19 | 3.5 (2.2,5.4) |
| Charlson score | | | | | | | | |
| 0 | 4428 | 13.4 (13.0,13.8) | 928 | 3.2 (3,3.5) | 2810 | 125.3 (120.7,130) | 928 | 3.2 (3.0,3.5) |
| 1–2 | 868 | 11.6 (10.9,12.4) | 244 | 3.6 (3.2,4.1) | 332 | 80.8 (72.5,89.9) | 244 | 3.6 (3.2,4.1) |
| 3+ | 223 | 13.1 (11.5,15.0) | 93 | 3.5 (2.9,4.3) | 49 | 59.1 (44.6,78.2) | 93 | 3.5 (2.9,4.3) |
| Intensity of testing per 100 days | | | | | | | | |
| <1 | 3550 | 9.2 (8.9,9.5) | 1010 | 2.8 (2.6,3) | 2429 | 97 (93.2,100.9) | 1010 | 2.8 (2.6,3.0) |

| | | | | | | | | |
|---|---|---|---|---|---|---|---|---|
| 1– <2 | 1472 | 53.2 (50.6,56.0) | 179 | 14.7 (12.7,17.1) | 619 | 336.5 (311.0,364.0) | 179 | 14.7 (12.7,17.1) |
| 2+ | 497 | 60.5 (55.5,66.1) | 76 | 12.7 (10.2,16) | 143 | 290.4 (246.5,342.1) | 76 | 12.7 (10.2,16.0) |
| Time since the last immunity conferring event | | | | | | | | |
| <100 days | 4570 | 12.2 (11.9,12.6) | 179 | 3.1 (2.7,3.6) | 3009 | 115.9 (111.9,120.1) | 179 | 3.1 (2.7,3.6) |
| 100-199 days | 949 | 19.7 (18.4,20.9) | 815 | 3.1 (2.9,3.3) | 182 | 128.2 (110.9,148.3) | 815 | 3.1 (2.9,3.3) |
| ≥200 days | | | 271 | 4.7 (4.2,5.3) | | 116.7 (112.7,120.8) | 271 | 4.7 (4.2,5.3) |
| No of immunity conferring events | | | | | | | | |
| 1 | 5491 | 13 (12.7,13.4) | 1254 | 3.3 (3.1,3.5) | 3189 | 36.6 (9.1,146.2) | 1254 | 3.3 (3.1,3.5) |
| ≥2 | 28 | 16.1 (11.1,23.3) | 11 | 7.1 (3.9,12.7) | 2 | | 11 | 7.1 (3.9,12.7) |

**Table S4. Factors associated with SARS-CoV-2 infection (adjusted HRs together with 95% CI), Estonia 2020–2022.**

| | **COHORT 1: NATURAL VS NO-IMM** | | **COHORT 2: HYBRID VS NATURAL** | | **COHORT 3: VACCINE VS NATURAL** | |
|---|---|---|---|---|---|---|
| | **Delta** | **Omicron** | **Delta** | **Omicron** | **Delta** | **Omicron** |
| | adjHR (95% CI) | adjHR (95% CI) | adjHR (95% CI) | adjHR (95% CI) | adjHR (95% CI) | adjHR (95% CI) |
| Exposed vs unexposed | 0.17*** (0.15,0.19) | 1.26*** (1.19,1.33) | 0.61*** (0.46,0.80) | 1.05 (0.93,1.19) | 4.90*** (4.48,5.36) | 1.13*** (1.06,1.21) |
| Sex | | | | | | |
| Female vs male | 1.21*** (1.14,1.29) | 1.18*** (1.12,1.25) | 1.25* (1.03,1.53) | 1.05 (0.97,1.14) | 1.03 (0.98,1.08) | 1.12*** (1.08,1.17) |
| Age groups | | | | | | |
| 50–64 vs <50 years | 0.72*** (0.67,0.78) | 0.67*** (0.63,0.72) | 0.62*** (0.48,0.80) | 0.68*** (0.62,0.75) | 0.82*** (0.78,0.87) | 0.68*** (0.64,0.71) |
| 65–79 vs <50 years | 0.50*** (0.44,0.57) | 0.37*** (0.33,0.42) | 0.72 (0.51,1.02) | 0.37*** (0.31,0.45) | 0.58*** (0.54,0.63) | 0.38*** (0.35,0.41) |
| 80+ vs <50 years | 0.48*** (0.40,0.57) | 0.19*** (0.15,0.25) | 0.83 (0.53,1.30) | 0.26*** (0.19,0.37) | 0.60*** (0.54,0.67) | 0.17*** (0.14,0.19) |
| Nationality | | | | | | |
| estonian vs else | 0.68*** (0.63,0.72) | 0.91*** (0.86,0.96) | 0.80* (0.65,0.98) | 0.97 (0.90,1.05) | 0.78*** (0.73,0.82) | 1.04 (0.99,1.08) |
| Education | | | | | | |
| higher vs below higher | 0.85*** (0.79,0.92) | 0.92* (0.87,0.98) | 0.73** (0.58,0.91) | 0.99 (0.91,1.07) | 1.03 (0.98,1.09) | 1.11*** (1.06,1.16) |
| Charlson score | | | | | | |
| 1–2 vs 0 | 1.12 (1.00,1.27) | 1.16** (1.05,1.29) | 1.54** (1.16,2.06) | 1.03 (0.89,1.18) | 1.12*** (1.05,1.20) | 0.98 (0.92,1.05) |
| 3+ vs 0 | 1.62*** (1.31,2.01) | 0.8 (0.62,1.04) | 1.38 (0.85,2.26) | 0.89 (0.66,1.21) | 1.26*** (1.12,1.42) | 0.80** (0.70,0.92) |
| Intensity of testing per 100 days | | | | | | |
| 1–<2 vs <1 | 3.21*** (2.98,3.46) | 2.80*** (2.62,2.98) | 4.57*** (3.65,5.71) | 2.39*** (2.16,2.63) | 5.04*** (4.75,5.34) | 2.37*** (2.22,2.53) |
| 2+ vs <1 | 3.21*** (2.86,3.61) | 2.98*** (2.69,3.31) | 6.11*** (4.67,7.99) | 2.55*** (2.22,2.94) | 6.15*** (5.63,6.72) | 2.29*** (2.06,2.54) |
| Time from last immunization (natural or vaccine) per month | 1.03*** (1.01,1.04) | 1.02** (1.00,1.03) | 1.09*** (1.05,1.14) | 1.01 (0.99,1.03) | 1.13*** (1.12,1.15) | 1.01** (1.00,1.02) |

adjHR – adjusted to all variables in table
* p<0.05, ** p<0.01, *** p<0.001

**Table S5. Factors associated with COVID-19 hospitalization (adjusted HRs together with 95% CI), Estonia 2020–2022**

| | Cohort 1: Natural vs no-imm | | Cohort 2: Hybrid vs natural | | Cohort 3: Vaccine vs natural | |
|---|---|---|---|---|---|---|
| | **Delta** adjHR (95% CI) | **Omicron** adjHR (95% CI) | **Delta** adjHR (95% CI) | **Omicron** adjHR (95% CI) | **Delta** adjHR (95% CI) | **Omicron** adjHR (95% CI) |
| Exposed vs unexposed | 0.05*** (0.03,0.11) | 0.10*** (0.04,0.26) | | 0.7 (0.04,12.70) | 7.19*** (4.02,12.84) | 2 (0.64,6.25) |
| Sex | | | | | | |
| Female vs male | 0.59*** (0.44,0.77) | 0.43** (0.26,0.72) | 2.42 (0.26,22.65) | 0.29 (0.05,1.88) | 0.58*** (0.43,0.79) | 0.59 (0.29,1.21) |
| Age groups | | | | | | |
| 50–64 vs <50 years | 3.13*** (2.08,4.70) | 4.84*** (2.08,11.26) | | 0.71 (0.06,8.97) | 2.20* (1.20,4.05) | 1.13 (0.33,3.84) |
| 65–79 vs <50 years | 5.82*** (3.81,8.90) | 8.82*** (3.71,20.96) | 0.7 (0.03,17.27) | 0 (0.00,.) | 3.50*** (1.93,6.34) | 1.61 (0.48,5.44) |
| 80+ vs <50 years | 9.29*** (5.83,14.80) | 15.71*** (6.22,39.65) | 1.41 (0.05,37.47) | 6.78 (0.52,88.77) | 5.95*** (3.19,11.08) | 7.13** (2.19,23.21) |
| Nationality | | | | | | |
| estonian vs else | 0.77 (0.58,1.01) | 1.15 (0.69,1.91) | 1.25 (0.19,8.40) | 3.05 (0.31,29.89) | 0.98 (0.68,1.40) | 1.77 (0.80,3.90) |
| Education | | | | | | |
| higher vs below higher | 1.15 (0.81,1.64) | 0.75 (0.37,1.53) | 0.91 (0.09,9.39) | 0.67 (0.07,6.32) | 0.58* (0.38,0.90) | 0.6 (0.21,1.75) |
| Charlson score | | | | | | |
| 1–2 vs 0 | 1.45 (1.00,2.12) | 2.20* (1.20,4.01) | 2.47 (0.12,50.20) | 6.74 (0.71,64.08) | 3.21*** (2.22,4.66) | 1.19 (0.42,3.33) |
| 3+ vs 0 | 3.43*** (2.19,5.36) | 2.57* (1.07,6.17) | 9.89 (0.44,224.57) | 0 (0.00,.) | 4.80*** (3.07,7.50) | 5.29*** (2.07,13.53) |
| Intensity of testing per 100 days | | | | | | |
| 1–<2 vs <1 | 2.73*** (1.86,4.00) | 3.34** (1.61,6.91) | 3.69 (0.32,42.47) | 5.09 (0.44,59.28) | 6.02*** (4.03,9.00) | 5.05** (1.64,15.52) |
| 2+ vs <1 | 3.34*** (2.00,5.57) | 6.47*** (2.83,14.80) | 11.00* (1.48,81.54) | 16.93** (2.26,126.86) | 12.22*** (7.88,18.96) | 10.52*** (3.54,31.24) |
| Time from last immunization (natural or vaccine) per month | 1.03 (0.97,1.10) | 1.03 (0.91,1.16) | 1.15 (0.85,1.55) | 1.09 (0.73,1.63) | 1.24*** (1.14,1.34) | 0.97 (0.78,1.21) |

adjHR – adjusted to all variables in table
* p<0.05, ** p<0.01, *** p<0.001
